# Class-Modeling of Septic Shock With Hyperdimensional Computing

**DOI:** 10.1101/2021.05.21.21257481

**Authors:** Neftali Watkinson, Tony Givargis, Victor Joe, Alexandru Nicolau, Alexander Veidenbaum

**Affiliations:** Donald Bren School of Information and Computer Sciences, University of California, Irvine Donald Bren Hall, 6210, Irvine, CA 92697; Regional Burn Center, UCI Medical Center, 101, The City Dr S, Orange, CA 92868

## Abstract

Sepsis arises when a patient’s immune system has an extreme reaction to an infection. This is followed by septic shock if damage to organ tissue is so extensive that it causes a total systemic failure. Early detection of septic shock among septic patients could save critical time for preparation and prevention treatment. Due to the high variance in symptoms and patient state before shock, it is challenging to create a protocol that would be effective across patients. However, since septic shock is an acute change in patient state, modeling patient stability could be more effective in detecting a condition that departs from it. In this paper we present a one-class classification approach to septic shock using hyperdimensional computing. We built various models that consider different contexts and can be adapted according to a target priority. Among septic patients, the models can detect septic shock accurately with 90% sensitivity and overall accuracy of 60% of the cases up to three hours before the onset of septic shock, with the ability to adjust predictions according to incoming data. Additionally, the models can be easily adapted to prioritize sensitivity (increase true positives) or specificity (decrease false positives).

## I. Introduction

Sometimes, sever infections can cause an extreme immunological response that causes severe tissue damage to organs and eventually, death. The term for referring to this condition is sepsis, and it is the leading cause of hospital deaths in the United States (1 in every 5 deaths) [1]. With early detection, sepsis can be managed with antibiotics (when the source of infection is bacterial). Sepsis survival rate is over 70% if properly diagnosed. If sepsis goes unnoticed, or the infection is resisting treatment, a patient will suffer multiple organ dysfunction that causes a systemic shutdown. This phase is called septic shock [2].

Current protocol [3] defines the clinical criteria for detecting septic shock as the presence of hypotension that won’t respond to fluid resuscitation, accompanied by associated tissue hypoperfusion, and requiring the use of vasopressors. There are different proposed guidelines regarding when vasopressor usage should begin – the earliest suggested starting time being simultaneous to fluid resuscitation in response to hypotension [4], [5].

Detecting the onset of sepsis depends on the protocol or scoring system observed by the hospital [6]. The four most common systems used are:

- SIRS (Systemic Inflammatory Response Syndrome): A simple binary scoring system with 4 variables related to: Temperature, blood pressure, white blood cell count, and heart rate [7]. If each value is outside a defined threshold then the score is incremented by 1.
- MEWS (Modified Early Warning Score):Comprises 6 vital signs and assigns a score from 0 to 3 to each one according to predetermined thresholds. A cumulative score of 4 or more triggers a call to a Rapid Response Team.
- SOFA (Sequential Organ-Failure Assessment): Previously known as the Sepsis-related Organ-Failure Assessment. Since 2016, SOFA is part of the official Sepsis-3 definition [8], that defines sepsis clinically as an acute increase of 2 or more in SOFA score. It uses a cumulative score over a number of vital signs (6 groups in SOFA’s case). The primary objective for SOFA is to identify organ failure.

When compared in effectiveness for diagnosing outcomes of septic patients, SIRS has a tendency to be oversensitive (high number of false positives) [9], and MEWS was not specifically designed for sepsis, hence, accuracy in identifying it is low [10]. While SOFA has higher sensitivity and specificity than SIRS or MEWS, it is considered to be the most burdensome and impractical for continuous monitoring scenarios. Quick-SOFA (or qSOFA) is a version that uses a shorter criteria (3 values) that requires a pre-established suspicion of sepsis, and it’s meant to be a monitoring tool in non-ICU (Intensive Care Unit) settings [6], [11]. Some works have tried to produce a predictive tool that would help in defining a protocol for septic shock [12], [13]. The effectiveness in modeling septic shock has been limited [14]. In this paper, we describe modeling of septic shock using a one-class classifier based on hypderdimensional (HD) computing. The model targets septic patients in an ICU setting.

The rest of the paper is organized as follows:

- **Methodology** describes our HD computing implementation.
- **Results** shows a comparison between models that used different inputs for the same population.
- In **Discussion** we justify why this approach works and compare it to industry standards.
- **Conclusion** summarizes our findings and future work.

## II. Methodology

The primary goal is to create a system that can predict septic shock before it happens. The target population is ICU patients with confirmed sepsis. We identify our targeted event as the time when a patient’s mean arterial pressure (MAP) goes below 65 mmHg (hypotension) and/or lactate levels are over 2 mmol/L (tissue hypoperfusion). These are the two main indicatives or pre-ambles of septic shock.

### A. Hyperdimensional computing

HD computing is computing paradigm inspired by cognitive science [15].It uses high dimensional vectors to encode information. The mathematical properties of these vectors can be exploited for efficient pattern recognition and modeling.

A general overview of the training algorithm for HD computing is described as follows:

1. Encode feature values into k-dimensional vectors. Within the feature domain, each value (or value range in the case of continuous data) corresponds to a unique vector. In the case of binary vectors, the number of 1s and 0s should be equal (partially dense) in order to exploit mathematical properties of orthogonality among feature vectors (orthogonal vectors are dissimilar [16]). This means that the distance between vectors should not be dependent upon the difference in density, but about the dimensional distance between them.
2. For each data point (also known as instance or subjects, i.e., the units of a dataset), combine all the feature vectors to form a new k-dimensional vector. Common operations for combining these feature involve adding them through arithmetic addition (for vectors of scalar values), multiplication (for polar vectors where every number is 1 or −1) or majority voting (for binary vector where each number is 1 or 0).

After populating the training space, for each new data point or query introduced for classification, inference unfolds as follows:

1. Encode the data point using the same methodology used in training to generate a k-dimensional vector. This vector is called the *query vector*.
2. Compute the distance between the query vector and all the training vectors or class vectors.
3. Output the predicted class as that of the closest training vector.

In this work, we focus on binary 10k-dimensional (10 thousand elements) vectors and Hamming distance for prediction. The main advantages of HD computing within this specific project are:

- Dimensionality is constant and adaptable, regardless of the number and/or magnitude of the features.
- There isn’t an iterative training phase like most traditional Machine Learning (ML) have.
- The encoding algorithm relies largely on two simple math operations: addition and multiplication.
- Correlation is captured during encoding, making it highly adaptable to different domains. New data points can be incorporated without having to rebuild the model.
- Because classification is dependent on proximity, we can implement a one-class classifier natively.

However, there are relevant weaknesses when compared to classical ML approaches. Two of the main weaknesses are:

- Classification performance is highly susceptible to how encoding is done. This impact can only be assessed at run-time, relying on trial and error.
- Depending on the application, encoding may require extensive domain knowledge to understand feature correlation. Preserving linearity or evaluating feature relevance are not part of the main algorithm and more traditional approaches are currently needed.

We chose HD computing because we believe that for this problem, the strengths outweigh the weaknesses. We base our encoding and feature extraction on domain knowledge from previous work in this area. Furthermore, we can design the application around the data we have available.

The first major decision regarding HD computing is on how to encode the features. We use linear and n-gram encoding, as described below.

#### 1) Linear encoding

There are two ways commonly used for encoding values: orthogonal encoding [17] and linear encoding [18]. Because our features are continuous values for which is important to preserve the pairwise magnitude and distance between each other, we decided on using linear encoding. The algorithm, as applied to this application, works as follows:

1. For each feature, identify the lowest, *min(V)*, and highest, *max(V)* values.
2. Generate a random 10k binary vector that are partially dense (has an equal amount of 1s and 0s). This will be our seed vector and used to represent every value equal or lesser than *min(V)*.
3. For all other values, flip an equal *x* number of 0 and 1 bits from the seed vector according to the following formula:

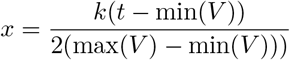 Where *k* is the dimensionality of the vectors, *t* is the target value, and *V* is all the values for a specific feature. Note that the range is doubled so that the highest value gets a vector orthogonal to the vector for the lowest value.

Using the formula for dynamic creation of vectors means that we don’t have to keep a vector dictionary in memory, adding efficiency to our application.

#### 2) n-gram encoding

Orthogonal encoding works well for discrete values that hold no relationship to each other. Linear encoding captures some of the relationship information from values within a feature. n-gram encoding is used for when the relationship is present between heavily correlated features, such as in the case of sequences. The name for this type of encoding comes from the n-gram model commonly used in information retrieval [19]. The general algorithm for binary vector n-gram encoding works as follows:

1. After encoding (using linear or orthogonal encoding) all the features, identify those that are part of a sequence and order them in descending order. For example, in a time sequence where each feature is a measurement for a different hour, then the feature with the oldest value goes first (feature 0).
2. Define the size of your n-gram. If three sequential features are joined then it will be a 3-gram.
3. Starting with the first feature (feature 0), permute (rotate the vectors by one bit) each vector *n-i* times where *i* is the feature index within the sequence and *n* the size of the n-gram.
4. Combine all the vectors in the n-gram using XOR. This will produce a new hypervector that will be semi-orthogonal to the individual vectors. This is key to represent the sequence as a new feature.

This encoding will generate new vectors that are almost geometrically orthogonal to the ones generated for the raw values, with similar sparsity. When building the final vectors that represent our data point (patient), we first add the raw vectors and the sequential values separately, using majority voting. This is necessary in order to keep sequential features from overtaking the final vector.

### B. One-Class Classification

OCC, also known as unary classification or class modeling, is a machine learning approach that focuses on identifying members of a single class without modeling an opposite (or negative) one [20]. The first published paper on OCC was by Moya, et al. [21], and it is a useful approach when a problem calls for finding a specific type of instance without the need or awareness of all other types or classes represented (for example, identifying spam documents among a body of documents from different domains) or when data about the complementary class is lacking or not available (a good use for this is detecting system failure using data that only represents a stable system). Figure 1 shows a representation of how an OCC model works. The model learns a single class, the *regular* class. The task is for the model to discriminate anomalies through elimination by inferring membership to the regular class. OCC is usually accompanied by a boundary threshold that determines how tolerant the model will be to instances far from the learned data.

**Fig. 1.**
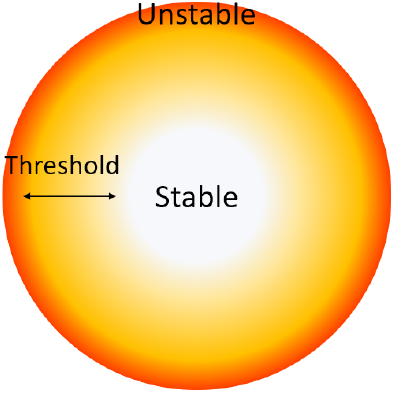
Graphical representation of OCC models applied to system stability. Data points far from the center are considered unstable according to the threshold chosen for the model.

Due to it’s nature, OCC is by design closer than multi-class classification to the neurological idea of core detection [22]. Even though plenty of research is available on multi-class classification and artificial neural networks (including deep learning algorithms), there are only a few examples where neural networks are applied to OCC [23]. At the time of this writing there was no method for building an artificial neural network or its derivatives (Deep Artificial Neural Network, Convolutions Neural Network, among others) for OCC. Most works modify and adapt multi-class networks to generate an OCC-like output [24]. OCC has been identified as a harder problem than multi-class classification mainly due to the challenge of tuning input parameters without looking at a sample of the opposite class [22].

Using OCC with HD computing hasn’t, to our knowledge, been explored yet. Our approach is similar to how OCC works within classical ML. In the case of HD computing, we encode the data that represents the stable or target class. For an unclassified instance, we measure its distance with all the members of the target class and choose the shortest one. The higher this distance is, the higher the probability that the instance is not a member of the target class. Alternatively, we can choose a distance threshold where everything with a distance equal or higher to it is considered a non-member.

Septic shock will progress differently, depending on variables such as which organs are failing. Septic shock can be considered as a system failure. Therefore, we can define our target or training class as stable or healthy patients, and the farther a patient is from that class the higher the probability of a system failure happening.

## III. Results

One of the most popular databases for health informatics research is MIMIC-III [25]. However, MIMIC-III uses an outdated definition of sepsis and its subsets. This has great impact in how the class is represented. Instead, we use eICU [26], a newer database from the same team behind MIMIC-III that uses the updated definition of sepsis.

Since eICU has data from patients within the ICU, we don’t have access to data from healthy patients. On the other hand, the problem we are trying to solve is to detect patients with confirmed sepsis that are at risk of going into shock. Due to data constraints, we will focus on bacterial infection related sepsis. We don’t consider fungal or virus related sepsis due to lack of samples. We also omit sepsis in burn and cancer patients due to different treatment interactions and symptom overlap between the two.

The general population criteria is: patients admitted into the ICU with confirmed sepsis (using Sepsis-3 criteria) who were not in shock when admitted, underwent antibiotic treatment, and have at least three hours worth of data. Using this criteria, We obtained data from 1237 patients before applying the data filters for the positive and negative classes. Table I displays the value distribution of three relevant features for each class.

**TABLE I.** Distribution for three relevant features within the positive (septic shock) and negative (not septic shock) classes. The value represents the average and the range (inside the parentheses).

| Feature | Positive | Negative |
| --- | --- | --- |
| Age | 63 (17-89) | 61 (18-89) |
| WBC | 11 (0.1-42) | 10 (0.6-46) |
| MAP | 83 (65-144) | 94 (65-147) |

### A. Positive class

Before trying the one-class approach, we built a baseline model that performs binary HD classification. Within the context of binary classification, the positive class is built with patients who, in addition to our population definition, underwent sepsis related hypotension (MAP<65 mmHG) and/or hypoperfusion (lactate>2 mmol/L), and were not being treated with vasopressors previous to this event (387 patients). The timestamp for this event is denominated as Hour Zero (H0).

### B. Negative class

The complementary, or negative class, are patients from the defined population who didn’t have a shock related event during their stay, were not treated with vasopressors, and were released from the hospital without dying (567 patients).

#### 1) Features

We consider two types of features:

- Sequential or periodic: These are values that change overtime and are available to build sequences. We consider 4 sequential features:
  – Mean Arterial Pressure (MAP)
  – Respiration Rate
  – Oxygen Saturation (SaO2)
  – Heartrate
- Static: These are features that don’t change within the observation window, we consider 6 features:
  – White Blood Cell Count (WBC)
  – Red Cell Distribution Width (RDW)
  – Blood Urea Nitrogen (BUN)
  – Creatinine
  – Sodium
  – Age

The criteria for feature selection is defined by availability of the data, and relevance within related work that point towards a correlation between the feature and sepsis/septic shock. For example, temperature is a feature that is highly correlated with sepsis and infection in general, but surprisingly, it isn’t widely available for all patients in the database.

For the positive class, sequential features are collected at 1, 2 and 3 hours before H0, considering the hourly average. For the negative class, collected features are from 1, 2 and 3 hours after sepsis is confirmed and antibiotic treatment has started, or after admittance, whichever is later.

#### 2) Synthetic data set

Since all the population in our data is from septic ICU patients, we don’t have a reliable baseline for our target class. Instead we created a synthetic data set with feature values that are considered as *healthy* by current medical standards. The feature values for the synthetic patients are randomly generated within a *healthy* range. Then the data is encoded using the same methodology used for the real patients.

### C. Binary Classification

The following models follow the classical HD computing algorithm for binary classification as presented in [15]. The purpose of these models is to understand the relationship between the positive and negative classes.

#### 1) Raw value encoding

We built three models that use the static features plus sequential features for each respective hour. Table II shows that there’s a slight increase in accuracy from the majority class (in this case, the negative class) for every hour. The last hour has the highest overall accuracy but all three models are similarly distributed.

**TABLE II.**
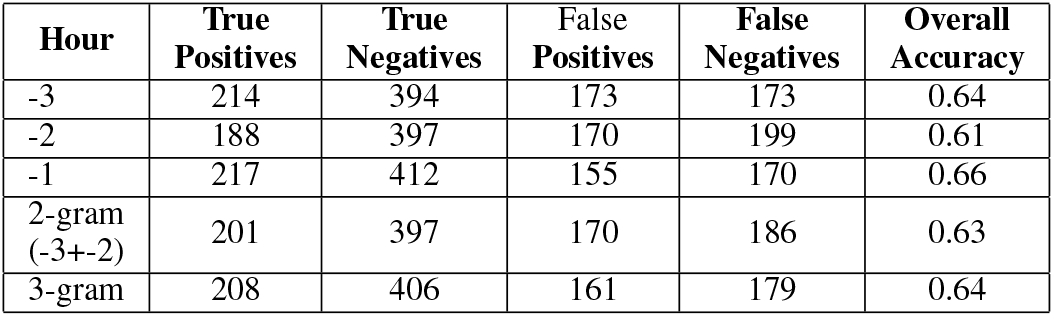
This table shows accuracy numbers for the HD computing model using binary classification. Hours are in the perspective of the positive class, −3 hours means 3 hours before hypotension is registered. For the negative class this would be the first hour of observation.

#### 2) n-gram encoding

Table II also shows how using n-gram encoding for sequential features performs. It is interesting that n-gram models seem to have similar accuracy numbers as the models that use raw values.

### D. One-class Model

In Table III, we describe the specificity at different sensitivity thresholds for the HD computing based one-class classifier (HD-OCC). This is a more descriptive perspective than reporting overall accuracy since the strength of this approach is in its adaptability of prioritizing the detection of positive cases or discarding false positives. While specificity seems low at high sensitivity thresholds, we must consider that the negative and positive classes are very similar before a patient goes into shock. Furthermore, replacing the synthetic dataset with real data or with a different generation approach to synthetic patients, could improve these numbers.

**TABLE III.** Specificity numbers for occ model using a synthetic dataset. The numbers for specificity represent specificity at sensitivity intervals of 0.9, 0.7 and 0.5 respectively

| Model | Specificity at sensitivity: |  |  |
| --- | --- | --- | --- |
|  | 0.9 | 0.7 | 0.5 |
| -3 hours | 0.16 | 0.45 | 0.61 |
| -2 hours | 0.16 | 0.47 | 0.66 |
| -1 hour | 0.19 | 0.51 | 0.66 |
| 2-gram (-3+-2) | 0.19 | 0.46 | 0.64 |
| 3-gram | 0.19 | 0.48 | 0.65 |

#### 1) Age multiplier

After further examination, we realized that age was having a detrimental effect on the model’s accuracy. A possible explanation is that while age is not necessarily an indicator of sepsis, it has a semi-linear correlation to poor outcomes. To represent the impact that age has with sepsis patients, we decided to not add age to the final vector through majority voting, but instead multiply the final Hamming distance by an age ratio (age divided by 40). This is in accordance with [27] that state that for patients older than 50 years old, the probability of septic shock increases. Therefore, distances for patients with an age close to 40 will stay overall the same, whereas higher ages will increase the final distance and lower ages will decrease them. While this approach remains to be validated, it did improve accuracy considerably.

Table IV shows the specificity numbers for the model with age as a multiplier. Most are an improvement over the previous model that encodes age as part of the patient vector.

To place this model into perspective, the best model for binary classification (−1 hour) has a sensitivity of 0.58 and specificity of 0.69. The one-class model has similar numbers at that threshold, but with the added benefit of being adjustable to prioritize sensitivity (reduce false negatives) or specificity (reduce false positives).

**TABLE IV.** Specificity numbers for OCC model multiplying distance by age. The numbers for specificity represent specificity at sensitivity intervals of 0.9, 0.7 and 0.5 respectively

| Model | Specificity at sensitivity: |  |  |
| --- | --- | --- | --- |
|  | 0.9 | 0.7 | 0.5 |
| -3 hours | 0.17 | 0.46 | 0.63 |
| -2 hours | 0.18 | 0.48 | 0.69 |
| -1 hour | 0.21 | 0.53 | 0.69 |
| 2-gram (-3+-2) | 0.19 | 0.46 | 0.66 |
| 3-gram | 0.19 | 0.50 | 0.68 |

## IV. Discussion

This paper primarily shows that HD computing has the potential of creating accurate models in both binary classification and OCC. At the time of writing, this is the first effort to apply HD computing to patient data.

However, the example application suffers from several limitations. A major obstacle is inconsistency within the data. There are features. such as temperature, that were missing from most patients in the dataset. For the few that had it, the value was measured outside the observation window, most often coming from a different hospital unit. Furthermore, this inconsistency has a negative effect on the granularity of the observation window. The data for the sequential features is continuous, however it was registered at different intervals. Because of this, we used hourly average instead, but this decision doesn’t allow us to use the full potential of n-gram encoding.

With more control over data input, such as using live data through an embedded solution, the models are bound to improve. N-gram encoding amplifies the distance to the synthetic dataset when there are abrupt changes in the information, but averaging sequential data has a smoothing effect. A similar conclusion was made by Van, et al. [28] after observing that increasing the time intervals had a negative effect on accuracy. For their model, increasing the intervals beyond 15 minutes per measurement meant a drop of classification accuracy down to 60% (from over 90% with one minute intervals). Part of our future work for this model is to gather data that provides sufficient granularity for predicting septic shock.

### 1) Feature cost

Feature cost refers to the effort (time, work, or in this specific case, intrusiveness of data collecting methods) needed to obtain it. An expensive or costly feature is one that requires more effort than the average or cheap features. Our model uses 4 features that are captured from the vital signs monitor.This monitor is commonly found as a bedside monitoring tool and once connected to the patient, commonly with supra-cutaneous diodes, measurements don’t require any intervention. In this context the features are considered to be cheap features. On the other hand, WBC, Creatinine, Sodium and RDW are more expensive because they require drawing blood from the patient and send it to a lab for analysis. As expensive as these features are, they are part of the same, standard protocol, blood screening. Since the backbone of the model are the cheap features, having new expensive data added can help the model’s accuracy but is not critical to the model’s performance.

On the other hand, SOFA uses features that require direct medical intervention or assessment and are mostly qualitative in nature. In Table V we compare the types of features each model uses,

**TABLE V.**
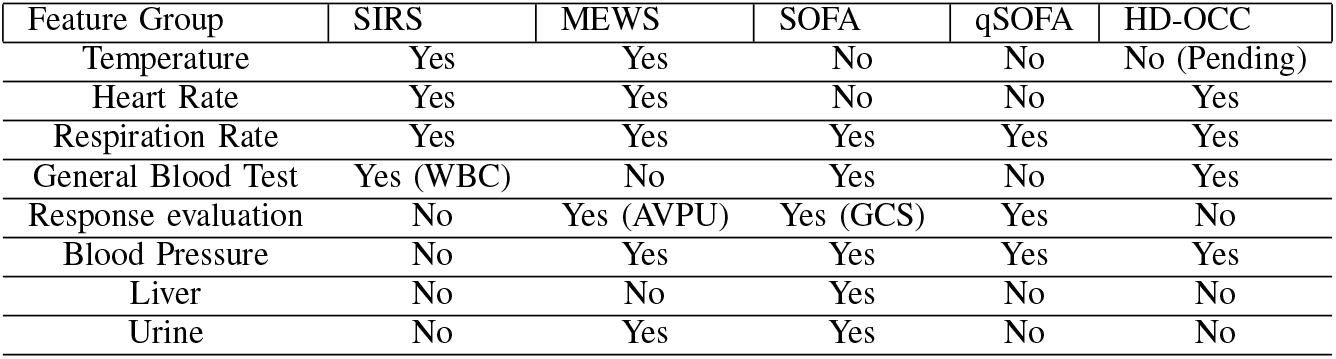
Feature table comparing our approach (HD-OCC) with industry standards. The first column is named Feature Group to categorize features that are closely related. Response evaluation encompasses different types of tests performed by medical staff to quantify the patient’s level of consciousness. GCS stands for Glasgow Comma Scale and AVPU stands for Unresponsive, Responds to Pain, Responds to Voice, and Alert.

| Feature Group | SIRS | MEWS | SOFA | qSOFA | HD-OCC |
| --- | --- | --- | --- | --- | --- |
| Temperature | Yes | Yes | No | No | No (Pending) |
| Heart Rate | Yes | Yes | No | No | Yes |
| Respiration Rate | Yes | Yes | Yes | Yes | Yes |
| General Blood Test | Yes (WBC) | No | Yes | No | Yes |
| Response evaluation | No | Yes (AVPU) | Yes (GCS) | Yes | No |
| Blood Pressure | No | Yes | Yes | Yes | Yes |
| Liver | No | No | Yes | No | No |
| Urine | No | Yes | Yes | No | No |

MEWS and SOFA are expensive models in terms of feature cost. SIRS and our model don’t rely on a qualitative assessment.

### A. Strengths and weaknesses of our approach

Unlike classical ML, our model allows us to use intuition and expertise to improve the model and make it robust. This model falls under a more specialized and tuned up approach that is adaptable to domain constraints.

Compared to industry standards, OCC combined with HD computing offers greater promise as an ever growing and always improving model. The more patients are treated, and more data is made available, the more opportunities for the model to improve and be finely tuned. This is a desirable property from an artificial intelligence approach.

This is not to say that there aren’t any drawbacks. Currently there isn’t a proven methodology for feature evaluation within HD computing. This means that new features need to be carefully evaluated with more conventional techniques since we have little insight on how they interact with the final class once added to the patient vector. HD computing requires domain knowledge and this is true also if the feature space is being expanded or the model is applied to a different target that is not septic shock.

Another issue is how descriptive the model is. HD computing is easy to understand mathematically. We can derive how features impact distance changes between patient vectors and gain some understanding on their relevance for predicting septic shock. In this sense it does a better job than most ML approaches at providing an explainable model. But we currently can’t provide a medically validated explanation to why a specific combination of feature values and threshold choice is considered close or far enough from the training vectors to be classified as septic shock, nor can we, if possible, translate the model into a non-dynamic criteria.

## V. Related work

While HD computing was proposed more than a decade ago, it is only in recent years that application oriented research started to appear. To our knowledge, there is no other project that has dealt with using HD computing to deal with patient data. In this section, we describe HD computing approaches inside the field of health informatics, but we also describe some of the current approaches to septic shock.

### A. HD computing in health informatics

One of the most prominent works in HD computing is that of Rahimi, et al [29] and [18] where data from non-invasive electrode’s is used to model brain activity and predict the subject’s intentions. They achieve a 5% improvement in accuracy over machine learning approaches. Similarly, in Imani, et al [30] they use HD computing for DNA modeling, achieving over 99% accuracy.

Though not within health informatics, Imani, et al [31] proposes an FPGA architecture that achieves over 5X performance when compared to other FPGA approaches. This is achieved by emulating an associative memory module with custom hardware configuration. Furthermore, Salamat, et al [32] describes an architecture that is up to 11 times more energy efficient than a GPU implementation. These findings are key factors for justifying the use of HD computing, specially when targeting constrained environments such as, in our case, next to a patient’s bed.

### B. Septic Shock modeling

The work of Giannini, et al. [13] solves a similar problem to the one presented in this chapter. However, they target patients outside the ICU. While the feature burden is equivalent to ours, they follow an old definition of sepsis. Furthermore, their model prioritizes specificity (over 90%) while sacrificing sensitivity (27%) without a way to modify the model.

The work of Kim, et al. [12] uses an ML approach to model septic shock. In this work they include chief patient complaint as a feature and encode it into numerical values. While their sensitivity (0.70) and specificity (0.90) are considerably higher than our approach, their study is retroactive to septic shock, with the observation window including patients that underwent shock already. Furthermore, their positive class is comprised of patients between 66 and 88 years old and the negative class age range is 51 to 79. Due to the little overlap between classes, they confirm age being a dominating feature in their model.

The challenge described in this chapter, however, is unique in the way the classes are defined. In the case of binary classification, the positive and negative classes are not linearly separable by any of the features and are similar to each other from the point of view of patient state. I target a critical observation window ensuring that patients have not gone through septic shock before making a prediction.

## VI. Conclusion

In this paper we described the application of a one-class classifier using HD computing for the prediction of septic shock among sepsis patients. The model performs consistently across use cases achieving up to 66% classification accuracy when balancing specificity and sensitivity but can be adjusted to prioritize one or the other. We argue that the application of this model doesn’t pose an extra burden on current hospital protocols and can provide an early alarm for preparation or prevention of septic shock. HD computing classification shows promise and allow for efficient models tailored for the problem we are trying to solve. In this case, HD computing enables us to use one-class classification for greater accuracy and flexibility to changing conditions when compared to binary classification.

## Data Availability

The data we used for our models is available through the resources we cite in the paper.

